# COVID-19 & Mental Health: Impact on Working people and Students

**DOI:** 10.1101/2021.08.05.21261663

**Authors:** Kshipra V. Moghe, Disha Kotecha, Manjusha Patil

**Author notes:** Corresponding Author: Disha Kotecha, Email ID.

## Abstract

A total of N=618 responses (16-60 years) were recorded to gauge the impact of COVID-19 socially, personally, and psychologically. Comparative results based on employment status, gender, and background were evaluated to identify the impact. While all the groups maintain having information about the pandemic and necessary safety protocols, there is an observable difference in the apprehension levels of financial and mental stability. Due to job security, employed people are less tense and better connected to their family, while unemployed people and students are more concerned with their productivity and quality of work. Students also display higher feelings of uncertainty and helplessness. A considerable number of people feel lonely and deserted during the pandemic. Such thoughts may leave a lasting effect if not tackled at the earliest. While an increase in awareness about mental health is observable, rural and unemployed people are less inclined to approach a professional. The significant difference COVID 19 has created between working people and students and based on gender and background, suggests that the preventive measures to avoid its lasting effects must be devised separately.

## Introduction

COVID-19 is a contagious disease that spreads via coughing, sneezing, or close contact. Patients can be symptomatic-experiencing high fever, body pain, breathlessness, pneumonia, or respiratory diseases within 2 to 14 days of incubation or asymptomatic-not showing any symptoms. People are advised to wear masks, maintain 3ft distance, and avoid public or crowded places until a vaccine or medicine is created (He et al., 2020) (Sharma & Nigam, 2020). Despite the precautionary measures, Coronavirus has affected approximately 213 countries with 92.3M total cases and 1.98M fatalities. Currently, India has 10.5M cases and 156K deaths as of 14 January 2021 (“Coronavirus Disease,” 2020). After the virus’s origin was found, China reported the first case in December 2019, where 11 out of 41 patients were severely ill and required immediate hospitalization (“Disease outbreak news,” 2020). In a short span, the virus had devastating effects on various countries like China, Italy, France, etc., following which it was declared a pandemic by the World Health Organization (WHO). In India, the first case was noted when a man with a travel history from Wuhan, China tested positive for Coronavirus in January 2020. As cases began to rise, the Indian Government declared a countrywide lockdown on 25 March 2020 to restrict its escalation. Stores, facilities, and businesses related to necessities like, medical facilities, groceries, pharmacies, etc. were authorized to operate.

From being a health concern, COVID-19 quickly widened to be an economic, social, and psychological nuisance. The reallocation of funds, shutting down of the global market, and halting of services and production created a scarcity of food and resources, imposing a financial and economic burden and, in turn, affecting the citizens (Ebrahim et al., 2020). On top of that, quarantine, self-isolation, and reduced social interaction due to lockdown and social distancing protocols give way to increased feelings of loneliness, distrust, and fear amongst citizens that can have a detrimental and irreversible effect on their mental health (Fiorillo & Gorwood, 2020). Front line workers like healthcare professionals and sanitation workers face higher physical and mental health challenges due to a shortage in protective equipment supply and lack of formal training in infection control, discrimination, social stigma, fear of life, and staying away from indefinite family period.

Constant uncertainty in terms of future, financial burden, striving to maintain a work-life balance, mainly those with children, and managing household chores are some issues the working adults as well as students face due to working or learning from home. Lockdown has hindered vacations, socialization activities, and breaks that are necessary for winding down during work. Dealing with these circumstances can cause emotional exhaustion and negative emotions, especially in the absence of assistance or support system. Hence, it is essential to connect with people virtually for emotional and social support (Ashida & Heaney, 2008).

As colleges and schools continue through online modes, the absence of face-to-face contact, monotony in routine, lack of facilities to enable proper learning, and dilemma about the future can have adverse mental effects. With a tendency to be vulnerable and develop symptoms of stress, anxiety and depressive thoughts (Mahmoud et al., 2012), those living in an abusive environment or living alone get exposed to prolonged stressors associated with anxiety, frustration, and depressive disorders (Bostan et al., 2020; Fiorillo & Gorwood, 2020). Avoidance behavior and a sedentary lifestyle can also develop due to change in sleep patterns, disturbance in or lack of routine, financial stress, social media pressure, or uncontrolled use of electronic devices. It is also imperative to regulate electronic devices to avoid overloading or false information from social media platforms (Jungmann & Witthöft, 2020). Dealing with estranged friendship, loss of loved ones, loneliness, and self-blaming can increase anxiety, depression, and tendencies of self-harm, alcoholism, substance abuse, or suicide amongst the populace (Matthews et al., 2018).

Research review of studies from countries like Turkey, Italy, China, Germany, and the USA showed the effect of COVID-19 on the general populace in terms of policies, healthcare, etc (Bostan et al., 2020; Jungmann & Witthöft, 2020; Zhang & Ma, 2020; X. Zhang et al., 2020). An early study from China focused on how COVID-19 has led to increased anxiety, PTSD, and depression among the student population (Liang et al., 2020). Studies from India are majorly based on the effect of COVID-19 on the healthcare professional and at-risk patients (Dubey et al., 2020). Based on the Indian population’s response to an online survey, a study focuses on attitude, anxiety, and perceived mental healthcare among the adult population (Roy et al., 2020), while in contrast, another focused on the pandemic’s impact on the student population socially, personally, and psychologically (Moghe et al., 2020). We aim to compare the impact of COVID-19 on the student and working adult population (which includes individuals currently employed and unemployed) of India so that specific intervention mechanisms for each category can be made.

## Methods and Material

In this study, online survey method was used for data collection. Incidental sampling was used by reaching out to the identified population of students and working professional via social media platforms, internal mailing systems, and formal and informal communication channels. Throughout the process, ethical constraint of confidentiality in data collection was followed. The survey questionnaire was designed by the authors which focused on four parameters: demographics, impact of COVID-19 on social, personal, and psychological wellbeing. The social impact included questions based on awareness about the pandemic, lockdown, and social distancing protocols, while personal impact covered responses to differences in daily routine, work productivity, financial apprehension, social media usage, the effect of relationships, and goals. The psychological impact section involved questions based on anxiety, stress, depression, suicidal thoughts, indulgence, addictive behavior, and perception about mental health. The social and personal impact section responses were assessed using a five-point Likert scale (Strongly Agree, Agree, Partially Agree, Disagree, and Strongly Disagree). In contrast, a ten-point rating (10% during COVID-19 to 100% during COVID-19) assessed the response to questions involving symptomology and impact on overall mental health. A total of N= 618 responses from various age groups. A detailed description of the demographic criteria is listed in Table 1.

**Table 1.** Demographic details of N=618 participants

|  | Category | No. of participants (n) |
| --- | --- | --- |
| Age | 16-25 | 389 |
|  | 25-45 | 127 |
|  | 45-60 | 83 |
|  | above 60 | 18 |
| Gender | Female | 162 |
|  | Male | 186 |
|  | Prefer not to say | 3 |
| Background | Urban | 280 |
|  | Rural | 71 |
| Family Type | Joint | 288 |
|  | Nuclear | 63 |
| Relationship Type | Single | 366 |
|  | Married / In a Relationship | 252 |
| Socio Economic Status | Working class | 8 |
|  | Lower- middle class | 50 |
|  | Middle class | 212 |
|  | Upper- middle class | 78 |
|  | Upper class | 3 |
| Work Status | Student | 361 |
|  | Work full time | 128 |
|  | Self-employed | 45 |
|  | Not employed | 52 |
|  | Retired | 19 |
| Family affected by COVID19 | No | 565 |
|  | Yes | 53 |

## Results

To gain insight into the necessary statistics and data analysis, Python’s Panda library was used while visualization was performed using the Seaborn library on Anaconda’s Jupyter Notebook. Comparison within groups belonging to different employment statuses, i.e., students in the age group of 16-25, employed individuals currently working part or full time, and non-employed, including people retired from their respective professions, performed using one-way ANOVA on IBM SPSS Statistic 19. The results from each category are summarized below:

1. **Social Impact:** The first section of the survey included questions based on social scenarios during the pandemic and the participant’s perception of it. The mean, variance, and standard deviation of these responses are stated in Table 2. Most of the respondents agree that they have sufficient information 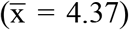, follow social distancing protocol 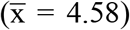, and the lockdown was a necessary step to tackle COVID-19 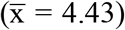 as depicted in the table. However, a considerable variation in their opinion on Unlock 1.0 (σ = 1.16) and eliminations of social barriers because of COVID-19 (σ = 1.23) can be observed. While some report an increase in mental health awareness, lockdown effectiveness, and news/ media channel as an information source, varying opinions also exist.

**Table 2.** Response to Social Impact of COVID-19
2. **Personal Impact:** The responses of participants to questions based on a five-point Likert scale were divided into three categories, namely, student (n=361), employed (n=186), and unemployed (n=71). These responses were further analysed using the one-way ANOVA and tabulated in Table 3. The significance factor (ɑ) chosen for comparison is p <= 0.05.

| Questions | Mean ($\bar{x}$) | Var ($\sigma^2$) | SD ($\sigma$) |
| --- | --- | --- | --- |
| Sufficient information | 4.37 | 0.41 | 0.64 |
| Practice social distancing | 4.58 | 0.32 | 0.57 |
| Lockdown has been a necessary step | 4.43 | 0.51 | 0.71 |
| Lockdown has proved to be effective | 3.85 | 1.00 | 1.00 |
| Unlock 1.0 introduced at right time | 3.10 | 1.30 | 1.16 |
| COVID-19 has eliminated social barriers | 3.02 | 1.51 | 1.23 |
| News/media were knowledgeable | 3.69 | 0.75 | 0.86 |
| Awareness of mental health increased | 3.55 | 0.81 | 0.90 |

**Table 3.** Response to Personal Impact of COVID-19 and comparison based on employment status There is a significant difference between employed, non - employed and students’ progress towards the work goal at the p<.05 level. The readings from the Mean scores suggest that a) the employed are better able to progress towards their goal compared to students and unemployed, b) postponing planned activities is lesser in the employed than unemployed whereas students postpone their activities the most, c) concern about career and finance is more in students than unemployed and least in employed. An analysis of developing positive relationship with family shows that employed seem to have improved compared to students and non - employed. Speaking of the physical health, students seem to have a significant increase in sleeping pattern than non - employed and employed; physical workout pattern is found to be not significant at the p<.05 level. For better insight, the data was further split based on gender, participants’ background, and current employment status. It was found that students (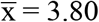, σ = 1.11) are spending more time on social media, than unemployed (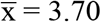, σ = 1.10) and employed (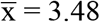, σ = 1.03); males (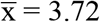, σ = 1.094) and participants from rural background (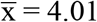, σ = 1.015) spend considerably more time on social media and are less conscious about their diet or food intake than the female (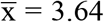, σ = 1.099) and urban (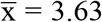, σ = 1.101) counterparts. Further, students (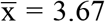, σ = 1.07) are more apprehensive about careers followed by the unemployed (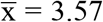, σ = 1.20) and the employed (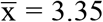, σ = 1.11); gender and background wise females (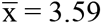, σ = 1.09) and rural (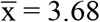, σ = 1.02) are more stressed about their careers. Also, while the employed (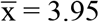, σ = 0.88) feels most positively closer to family as opposed to the unemployed (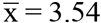, σ = 1.02) who feel least close; background wise the urban (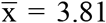, σ = 0.97) are found to feel closer to their families as compared to others.
3. **Psychological Impact:** These questions were based on a ten-point scale to understand the effect COVID-19 on the public psychologically. The responses were divided based on employment status (student (n=361), employed (n=186), and non-employed (n=71), after which one-way ANOVA was performed on the data at p<= 0.05 level of significance (Table 4). An analysis suggests an overall increase in unpleasant and painful feeling where a) students appear to have more unpleasant feelings than the employed and non – employed, b) significant increase in sense of helplessness and uncertainty which is more in students compared to the other two, c) significant difference between irritability, restlessness, and outbursts, with more effects in students compared to employed and non-employed. Considering health concerns and indulgence in intoxicants the difference is not significant. However, students seem to have significantly high indulgence in online activities compared to non-employed and employed hence a significant difference is observed in online indulgence.

| Employment Type | Mean ($\bar{x}$) | SD ($\sigma$) | f | sig.($\alpha=0.05$) |
| --- | --- | --- | --- | --- |
| Progress towards work goals |  |  |  |  |
| Students | 3.53 | .952 | 9.044 | .000 |
| Employed | 3.83 | .825 |  |  |
| Non – Employed | 3.39 | .933 |  |  |
| Total | 3.60 | .925 |  |  |
| Postpone Planned activities |  |  |  |  |
| Students | 3.57 | 1.068 | 6.037 | .000 |
| Employed | 3.24 | 1.019 |  |  |
| Non - Employed | 3.35 | 1.097 |  |  |
| Total | 3.44 | 1.066 |  |  |
| Apprehensive about career and finance |  |  |  |  |
| Students | 3.67 | 1.077 | 5.121 | .006 |
| Employed | 3.35 | 1.112 |  |  |
| Non - Employed | 3.58 | 1.203 |  |  |
| Total | 3.57 | 1.110 |  |  |
| Apprehensive about quality of work |  |  |  |  |
| Students | 3.56 | .923 | 11.118 | .000 |
| Employed | 3.20 | 1.019 |  |  |
| Non - Employed | 3.15 | .995 |  |  |
| Total | 3.41 | .977 |  |  |
| Positive relationship with family |  |  |  |  |
| Students | 3.75 | 1.001 | 5.183 | .006 |
| Employed | 3.96 | .887 |  |  |
| Non - Employed | 3.55 | 1.025 |  |  |
| Total | 3.79 | .978 |  |  |
| Increase in sleeping pattern |  |  |  |  |
| Students | 4.02 | .809 | 11.249 | .000 |
| Employed | 3.70 | .760 |  |  |
| Non - Employed | 3.77 | .687 |  |  |
| Total | 3.90 | .794 |  |  |
| Increase in workout |  |  |  |  |
| Students | 3.13 | 1.374 | 1.087 | .338 |
| Employed | 3.29 | 1.134 |  |  |
| Non - Employed | 3.27 | 1.172 |  |  |
| Total | 3.19 | 1.284 |  |  |

**Table 4.** Response to Psychological Impact of COVID-19 and comparison based on employment status Further, Figure 1a and 1b show difference in the responses based on gender and background, which suggests that participants from the rural background score high on almost all aspects compared to those from urban background, although the difference is negligible for increased intoxication intake and feeling helpless and uncertain. On the other hand, males’ scores vary from the female counterpart, showing a higher score only for increased online activities and an increase in intoxicants. The highest mean for participants from both male and rural is for increased online presence, deviating from the general norm.

| Employment Type | Mean ($\bar{x}$) | SD ($\sigma$) | f | sig.($\alpha=0.05$) |
| --- | --- | --- | --- | --- |
| Overall increase in unpleasant or painful feelings |  |  |  |  |
| Students | 5.45 | 2.959 | 7.154 | .001 |
| Employed | 4.58 | 2.780 |  |  |
| Non – Employed | 4.48 | 3.089 |  |  |
| Total | 5.07 | 2.950 |  |  |
| Overall increasing sense of helplessness and uncertainty |  |  |  |  |
| Students | 5.32 | 3.018 | 5.344 | .005 |
| Employed | 4.54 | 2.917 |  |  |
| Non - Employed | 4.46 | 3.175 |  |  |
| Total | 4.99 | 3.027 |  |  |
| Overall increase in irritability, restlessness, and outbursts |  |  |  |  |
| Students | 5.64 | 2.960 | 15.637 | .000 |
| Employed | 4.32 | 2.899 |  |  |
| Non - Employed | 4.25 | 2.902 |  |  |
| Total | 5.09 | 3.004 |  |  |
| Increase in overall health concerns |  |  |  |  |
| Students | 5.24 | 3.023 |  |  |
| Employed | 4.83 | 2.826 | 1.768 | .172 |
| Non - Employed | 4.69 | 3.017 |  |  |
| Total | 5.06 | 2.968 |  |  |
| Increase in overall indulgence intoxicants |  |  |  |  |
| Students | 1.40 | 1.502 | .313 | .731 |
| Employed | 1.49 | 1.438 |  |  |
| Non - Employed | 1.37 | 1.570 |  |  |
| Total | 1.42 | 1.490 |  |  |
| Increase in online indulgence |  |  |  |  |
| Students | 6.27 | 3.010 | 34.452 | .000 |
| Employed | 4.19 | 2.956 |  |  |
| Non - Employed | 4.38 | 3.109 |  |  |
| Total | 5.43 | 3.164 |  |  |

**Figure 1.**
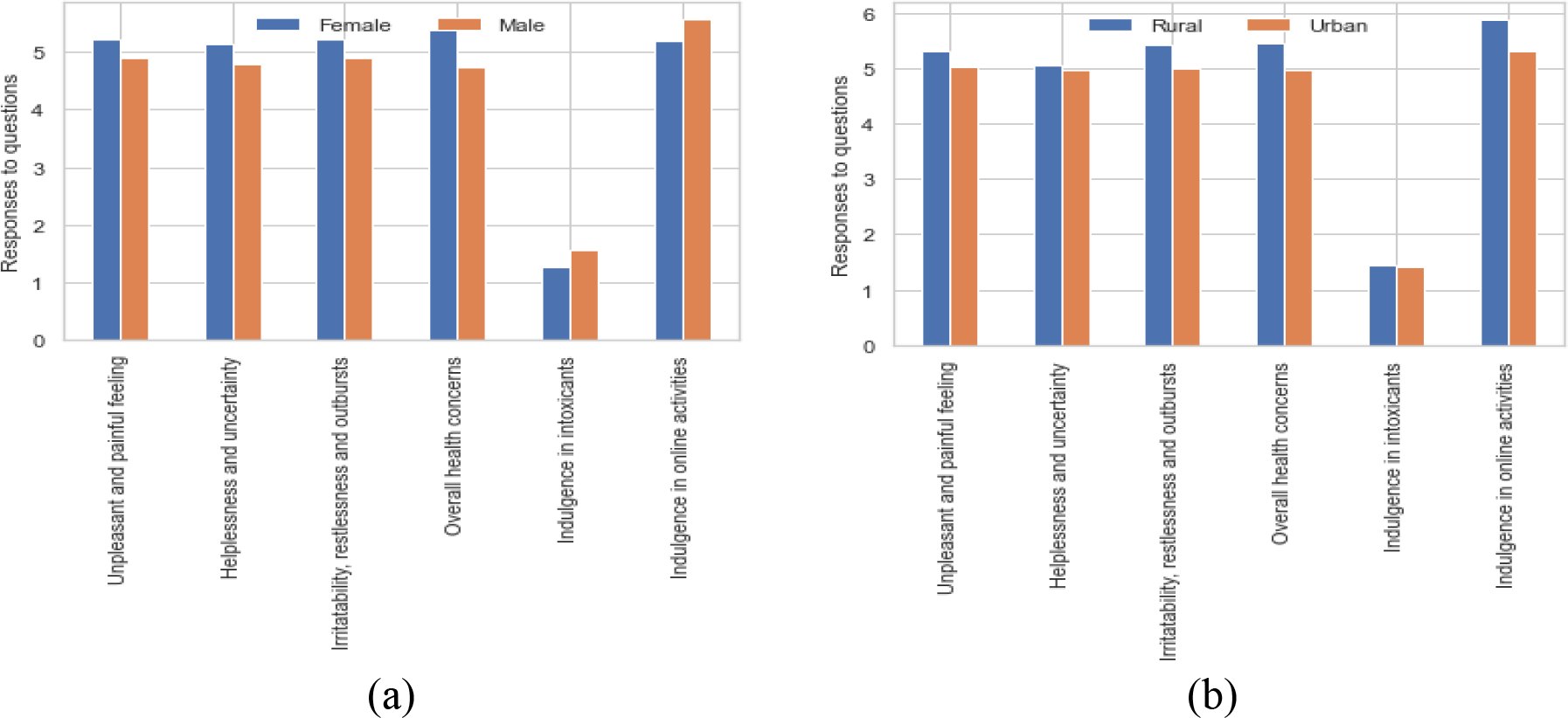
Bar graph representation of comparative statistic for the response to questions, (a) Based on Gender: Female/Male; (b) Based on Background: Rural/Urban Table 5 below depicts responses towards symptomology of stress, depression, anxiety, and suicidal thoughts based on a five-point Likert scale. Participants have majorly reported overthinking, feeling sad or empty, fatigue or restlessness, social withdrawal, or self-harm, and crying spells. Although the mean for other symptoms is comparatively lesser, it cannot be outright neglected. This data can be used for further focused intervention.

**Table 5.** Response to question based on symptoms of stress, anxiety, depression, and suicidal tendencies Self-perception, an important aspect of mental health, was also assessed (Table 6) and revealed that most of the participants across all the sections reported of becoming more practical during the pandemic, followed by becoming more self-aware and enjoying the solitude. Females, students and the unemployed also reported of becoming more complicated and uneasy during the quarantine. Furthermore, the highest number of students and females report feeling lonely or unwanted compared to groups in their respective categories.

| Questions | Mean ($\bar{x}$) | Var ($\sigma^2$) | SD ($\sigma$) |
| --- | --- | --- | --- |
| Rapid heart rate/ sweating/ pain/ chills/ hot flashes | 1.54 | 0.65 | 0.81 |
| Trembling/ shaking/ agitation/ throat dryness | 1.46 | 0.57 | 0.75 |
| Over thinking/ going in loops/ random thoughts | 2.57 | 1.81 | 1.34 |
| Crying spells/ losing control/ impending danger | 2.04 | 1.46 | 1.21 |
| Decrease in appetite/ water intake/ sleep/ sex drive | 1.85 | 1.20 | 1.09 |
| Mistrust/ preoccupation/ health alarm/ inability to relax | 1.83 | 1.12 | 1.05 |
| Feeling fatigued/ anxious/ restless/ imagining the worst | 2.07 | 1.50 | 1.22 |
| Feeling sad/ empty/ helpless/ guilty/ irritable | 2.22 | 1.69 | 1.30 |

**Table 6.** Comparison based on gender and employment status: Effect on self/ life perception during COVID-19 (in %) The bar graphs represented in Figure 2 (a) and (b) are comparative responses of participants’ willingness to contact mental health professionals and whether living with family has positively affected them. These questions were Yes/ No type, and the results represent a positive response from the participants. Most participants from rural and unemployed background are unwilling to consult a professional, along with males and people from joint families being less inclined towards the idea. The students and the female population reveal the highest positive response, although it is still less than 70%. Most of the people in each category (between 80% and 50%) are positively affected by living with family.

| Questions | Female | Male | Students | Employed | Unemployed |
| --- | --- | --- | --- | --- | --- |
| Religious/ Close to God/<br>Ritualistic | 5.82 | 6.23 | 4.71 | 6.45 | 11.26 |
| Spiritual/ Existential/<br>Introspective | 8.56 | 7.78 | 6.64 | 8.06 | 15.49 |
| Empathetic/ Compassionate/<br>Forgiving | 8.22 | 7.47 | 7.20 | 11.29 | 2.81 |
| Practical/ Realistic/ Prepared | 31.16 | 36.76 | 31.30 | 40.32 | 32.39 |
| Impatient/ Uneasy/ Difficult | 9.59 | 7.48 | 9.92 | 5.91 | 9.85 |
| Resilient/ Optimistic/ Grateful | 7.53 | 7.16 | 7.20 | 7.52 | 5.63 |
| Aggressive/ Resentful/<br>Jealous | 1.02 | 1.87 | 1.38 | 2.15 | 0.0 |

**Figure 2.**
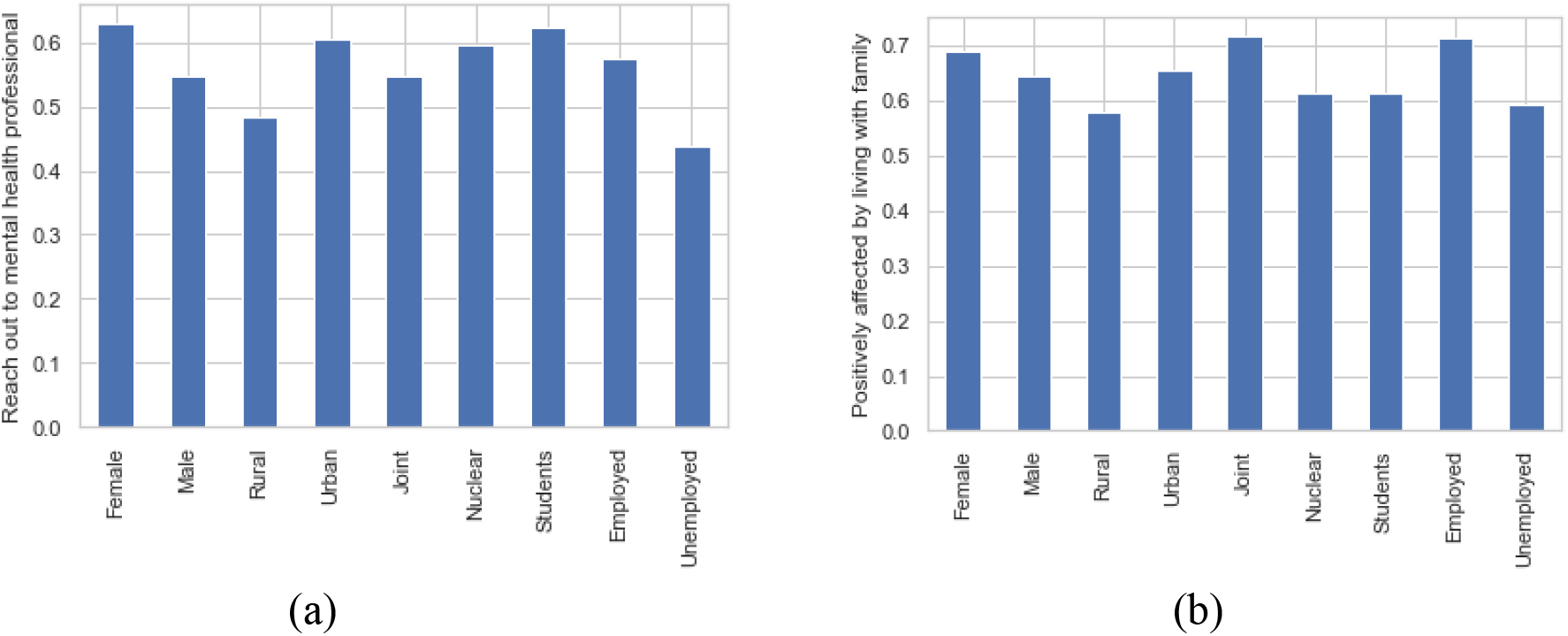
Bar graph representation of comparative statistic, (a) Reach out mental health professional (b) Positively affected by living with family

## Discussion

On average, the general population is aware of and abiding by the recommended safety measure of COVID-19. However, a lack of consensus on the effectiveness of Unlock 1.0 is seen which might be due to the ever-rising cases and usefulness of new and media channels. The employed seems to be faring better than their counterparts, which can be attributed to financial stability that comes with most jobs. Employed people are concerned about the uncertainty about the future and the economy, but they feel optimistic about the work from home option which might be a positive change from the routine and everyday stresses of traveling to work. While students report better progress in their goals, their concern about the quality of work, career, finances, and procrastination is more than the unemployed due to disruption of general routine, adjustment to the virtual learning, and ambiguity in the opportunities about getting a job market or higher studies. 63% of participants who have created a routine or timetable can progress towards their work goals better, while without a routine, only 20% seem to achieve their work goals. Also, creating a schedule that involves a workout, might help people adjust and handle the situation better, socially, physically, and mentally. Furthermore, although students appear to be more apprehensive about careers, their time on social media has increased, indicating a negative coping mechanism.

Results indicate higher mean values for students getting psychologically affected by COVID-19 compared to the employed and unemployed. Mental health and psychological wellbeing have been a relatively less discussed topic in India, and acceptance of related issues comes with challenges, especially with the older generations. This lack of acceptance and reluctance to acknowledge may be considered one of the explanations for the results and thus, may be taken at face value (Gruebner et al., 2017). Similarly, socially desirable responses might be a possible reason for the results indicating lesser indulgence of intoxicants over the actual scenario. This is in clear contradiction to what studies indicate about the increased consumption of intoxicants such as alcohol during the pandemic (Alpers, et.al., 2021; Bollen, et. al., 2021).

Results show that contrary to the popular opinion of lack of resources in the rural areas, the participants seem to be spending more time on social media, indicating the reach of internet services in the remote parts of the country. With the changing times and perceptions about gender roles, the results also indicate a shift in the increased concerns about careers by females; this is a welcome deviation from the general norm in Indian society where men are endowed to be the primary breadwinner in a family (Smojver-Ažić & Bezinović, 2011; Wardle et al., 2004).

COVID-19 has led to an increase in overall health concerns for everyone, irrespective of age. Results indicate an increase concern about health among those from the rural background, which may be due to the absence of advanced medical facilities and perceived fear of catching the virus. Results also indicate that females seem to be in touch with their emotions and express more as compared to males and more in touch with their feelings or express their emotions better. While this is quite expected, it also indicates that the pandemic might be more challenging for the males which can be further examined with a different study.

Further, results indicating a lower response on observed or perceived symptoms is also subject to being challenged since despite the number is meagre, it raises concern about the psychological wellbeing and a need for professional help to avoid prolonged damage. So is the case with results in Table 7, especially for those who report feeling lonely or becoming more jealous. It can leave a long-lasting impact, affecting them socially and mentally. Having said this, it must be noted that while 34% of participants consider that COVID-19 has made them more practical, realistic, and prepared, 17% have become more self-aware and are enjoying solitude, indicating that people are learning to adapt and adjust to make the best of the present situation.

The need for mental health awareness cannot be denied, however, the Indian society has its unique ways to dealing with less discussed issues-collectivistic culture in rural areas, joint family systems, and the support and guidance available from the older generations’ wisdom thus, appears to be positively helping the participants in the study. 68% of the sample feels that their most significant support has been their families, whereas 14% found friends or peers to be the most significant support. Most of the participants report of spending an average of 30-60 minutes with their families during the pandemic, whereas only 9% do not connect, emphasizing the role of relationships in times of adversities. Most of the students have responded to living with family as helpful, although not being able to socialize is an undesired outcome. And those considering self-support (3%) find their will power, self-confidence, and mental strength as factors for coping. Even though life appears to be monotonous, general positivity regarding increased time for self–introspection and more time for family and partner during lockdown is a welcome sign, evident from the results.

## Conclusion

The entire population is suffering from the repercussions of COVID-19. As the pandemic is far from being over, various coping mechanisms, not all of which are positive, have been adopted by people to adjust and continue living through it. For example, electronic devices and social media have increased drastically, causing people to become more withdrawn. The rise in worry regarding prospects of job and career, feelings of depression amongst students and unemployed people can cause lasting effects on the individuals. There is also a change in perspective concerning family and friends’ role as they have become the system of support. While employed people seem to be faring better in terms of work, they are more psychologically impacted by the pandemic than the non-employed population.

Similarly, while the need for mental health awareness is acknowledged by most of the population, their willingness to approach a professional is still low. It highlights the prevalence of stigma around mental and psychological health. It is deleterious considering the accounts of death and long-lasting results the COVID-19 can leave on individuals. Acknowledging the participants’ varying results and the need for positive coping mechanisms, individualistic intervention mechanisms should be devised.

## Limitations & Further Scope

As the sample size used for the study is limited to a single state, it is not a representation of the country-wide data and cannot be used for broader generalization. The statistical methods used here are limited, and for a better outcome, more complex algorithms can be adopted. This study aimed to find the correlation in the data between the working-class and students affected by COVID-19. A study carried out on a larger scale with additional variables can add more value to the data gathered during the times of COVID-19

## Data Availability

The data was collected from the consenting participants via online survey.

